# Short-term analysis and long-term predictions for the COVID-19 epidemic in a seasonality regime: the Italian case

**DOI:** 10.1101/2020.07.15.20154500

**Authors:** Giulia Simoni, Anna Fochesato, Federico Reali, Giulia Giordano, Enrico Domenici, Luca Marchetti

## Abstract

As of July 14^th^, COVID-19 has caused in Italy 34.984 deaths and 243.344 infection cases. Strict lockdown policies were necessary to contain the first outbreak wave and prevent the Italian healthcare system from being overwhelmed by patients requiring intensive care. After the progressive reopening, predicting how the epidemic situation will evolve is urgent and fundamental to control any future outbreak and prevent a second wave. We defined a time-varying optimization procedure to repeatedly calibrate the SIDARTHE model^1^ with data up to June 24^th^. The computed parameter distributions allow us to robustly analyse how the epidemic situation evolved and outline possible future scenarios. Assuming a seasonal regime for COVID-19, we tested different lockdown policies. Our results suggest that an intermittent lockdown where six “open days” are allowed every other week may prevent a resurgent exponential outbreak and, at the same time, ease the societal burden of an extensive lockdown.

## MAIN

The new strain of coronavirus SARS-CoV-2, causing a severe and potentially fatal respiratory syndrome named COVID-19, was initially identified in the Hubei province of China in the late 2019 and rapidly spread out worldwide, forcing the World Health Organization (WHO) to declare the pandemic alert on March 11^th^ 2020^2^. Most governments have established tight policies to slow down the spread of COVID-19, which has caused 548.211 deaths on July 8^th^. In Italy, one of the first and most affected Western countries, the interventions shifted from initial social distancing measures to a drastic nation-wide lockdown that started on March 11^th^ 2020, and lasted until May 4^th^. A massive swab campaign was set up to detect and isolate infected people, symptomatic and later also asymptomatic. From the beginning of May, more than two months after the Italian outbreak on February 20^th^ 2020, the situation is becoming under control, with a steady decrease of the number of new confirmed cases as well as hospitalised and Intensive Care Units (ICUs) patients. Such encouraging data allowed a cautious relaxation of the restrictive measures: gradual reopening of economic activities and free circulation of people. In this context, the ability to monitor and predict the disease progress is crucial to guide policymakers in deciding how to prevent and contrast possible future outbreaks.

Mathematical models offer a data-driven and quantitative understanding of the disease, allowing for probabilistic insights and future scenario predictions. Many researchers have proposed models for COVID-19^3–6^, building upon the common SIR-SEIR models for human-to-human transmission to describe the current epidemic. In this paper, we analyse the different phases of the spread of the disease in Italy using the recent SIDARTHE model^1^, which captures the population granularity by distinguishing between detected and undetected infected subjects as well as between asymptomatic and symptomatic infected subjects. We include as Extended Figure 1 a graphical representation of the model, described in Methods.

**Figure 1:**
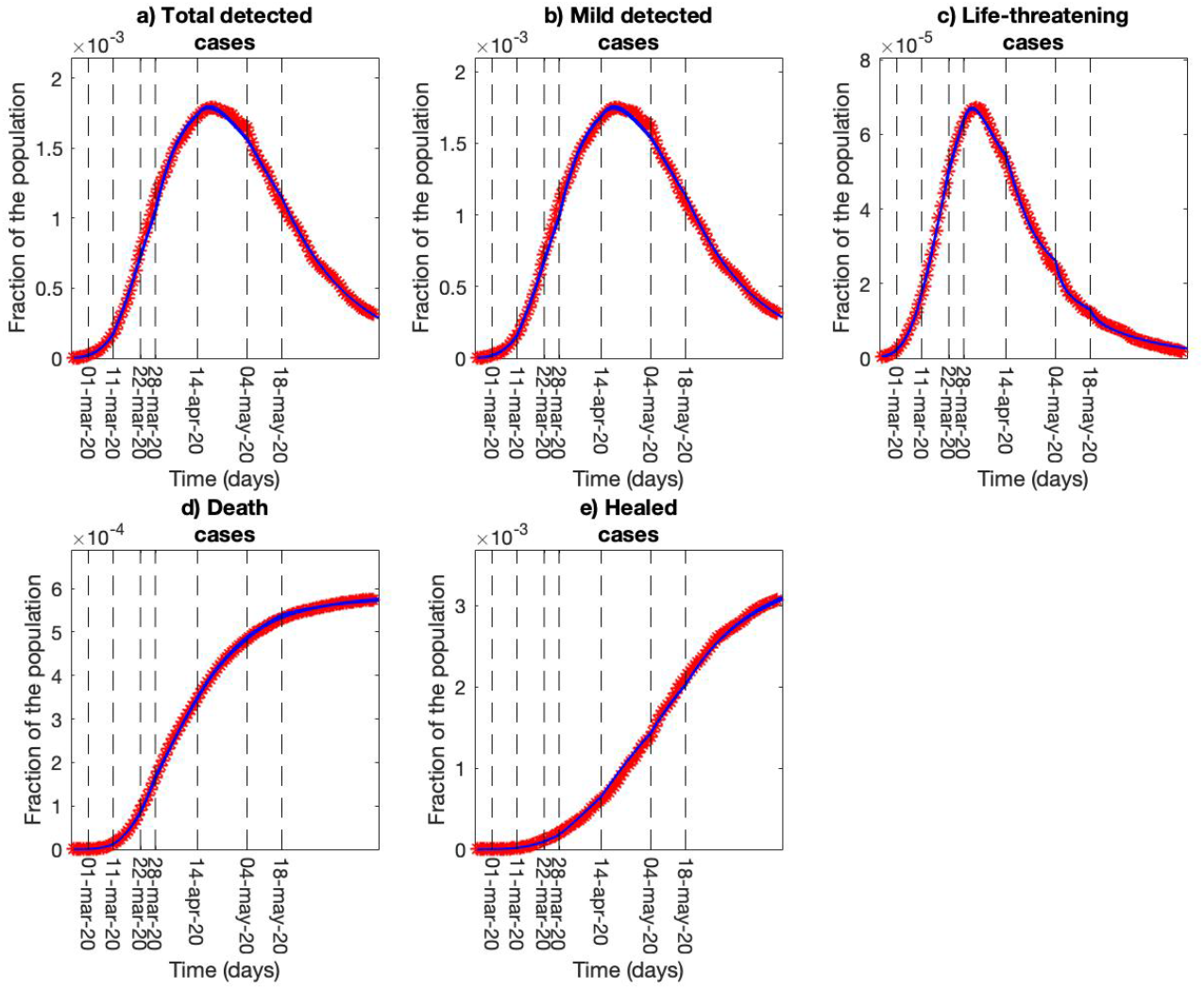
Model calibrations results. Short-term epidemic evolution obtained by repeated model calibrations on the basis of the public available Italian data. a) Daily evolution of the total detected cases computed as the sum of asymptomatic, symptomatic and life-threatening cases. b) Daily evolution of the mild detected cases computed as the sum of asymptomatic and symptomatic cases. c) Daily evolution of the life-threatening cases admitted to ICUs. d) Cumulative distribution of deaths. e) Cumulative distribution of the healed-detected cases. All the panels represent the data with red asterisks and the 100 model dynamics with blue lines. Both the data and the dynamics are normalized as fractions of the total Italian population. Dashed vertical lines represent a change in the social distancing or swab policies and a corresponding update of the model parameters.

We used a global optimization algorithm^7,8^ to estimate a subset of the model parameters on the basis of the data provided by the Italian Protezione Civile about detected cases, recovered, and deaths. We considered the period from February 24^th^ to June 24^th^ with an updating fitting strategy that reflects the shift in the adopted countermeasures, from full lockdown (Phase I), to partial restrictions (Phase II), to COVID-19-aware reopening (Phase III). The National Decrees of March 1^st^, 11^th^, 22^nd^, April 10^th^, May 4^th^, 18^th^, and the new swab policy of March 28^th^ have been used as critical events to mark the updates. We computed 100 repeated model calibrations to provide a robust distribution of each parameter estimate, thus relying on stable model predictions. More details about the time-varying calibration procedure can be found in Methods.

Figure 1 shows the short-term evolution of the model dynamics compared with the data used for the calibrations. In Extended Figure 2, the estimated model parameters show a decreasing trend for infection rates and an increasing trend for the recovery rates during the considered period. This reflects the efficacy of the restrictive measures in slowing down the virus transmission and the acquired experience in treating patients. In accordance with the model parameter trend, we observe a smooth decreasing curve for R0 that starts from a maximum value around 6 for February 24^th^ and crosses the critic value of 1 at the end of March^9^ (Figure 2a). The quartile curves highlight more confidence in the tail of the data period when the value of R0 became less than 1, suggesting a reliable estimation in the sensitive domain (0,1).

**Figure 2:**
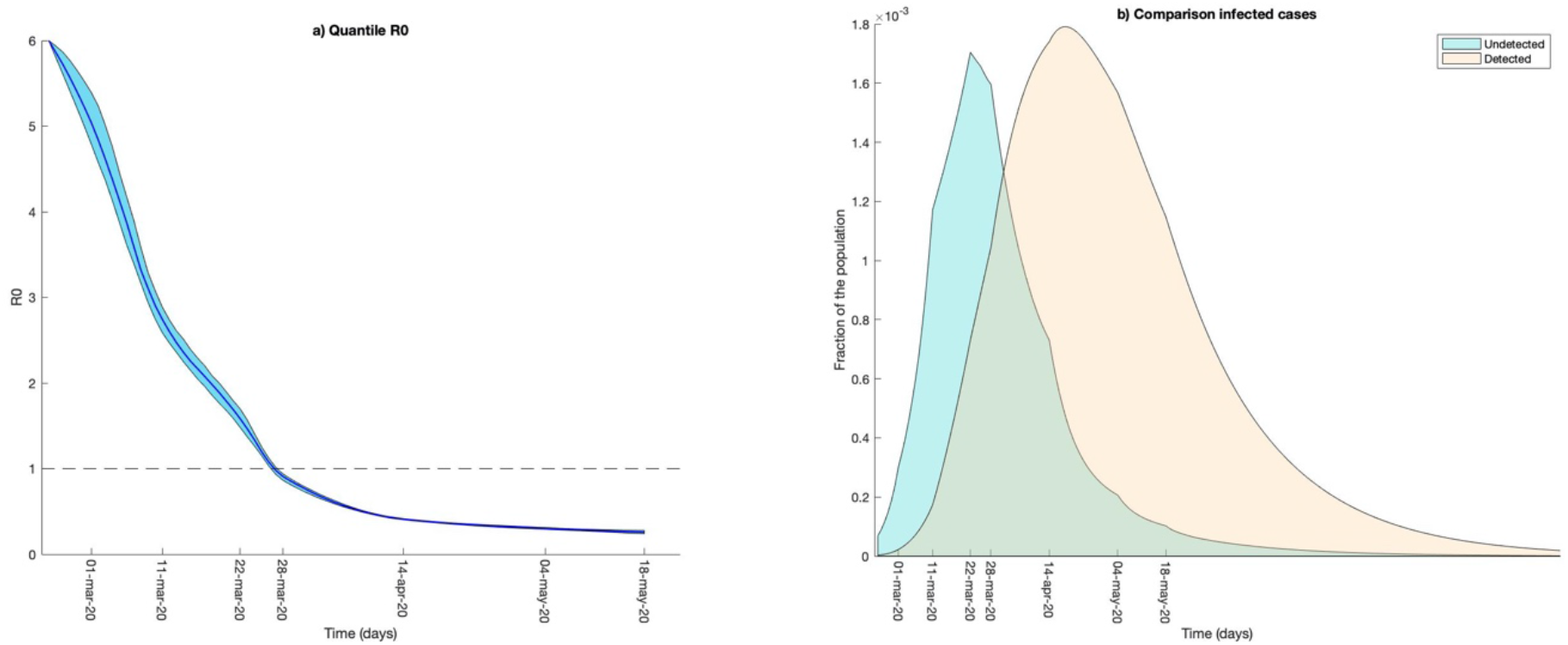
R0 and infected cases. From the repeated model calibrations, we computed the distribution of the model parameters and the corresponding distribution for R0. Besides, since the model accounts also for undetected individuals, we can compare the distribution of detected and undetected cases along time. a) R0 distribution. The light blue shade represents the values between the first and the third quartiles computed with the estimate distribution of model parameters obtained from the repeated model calibrations. The dark blue line represents the second quartile of the distribution. b) Comparison between undetected cases (variables I and A in the model) and detected cases (variables D, R and T in the model).

Combining the granularity of the SIDARTHE model with our fitting protocol allows us to estimate the undetected cases, who would need to be traced and tested in order to control the spread of the epidemic ^10,11^. In agreement with the estimation of Pedersen et al.^12^, we observed an initial ratio between undetected and detected cases of around 10:1 (Figure 2b). This result provides a possible quantitative explanation for the Italian exponential spread of COVID-19 in late February and early March due to the undetected circulation of the virus. In addition, the model suggests that the effects of the social distancing measures and the swab campaign led to a decrease of this ratio, up to a reversal reflecting an increasing control and management of the epidemic. Indeed, to avoid a second outbreak, it is crucial to identify and isolate the suspected infection cases with an efficacy testing policy^13–15^. In this way, we could be able to avoid an uncontrolled spread of the virus, thus reducing the risk of a new outbreak.

Figure 2b highlights how the number of undetected cases is prevailing in the early stage of the epidemic curve, suggesting that a relevant number of asymptomatic and undetected patients were present in Italy^16–18^, as well as in the rest of Europe, even before the implementation of the COVID-19 surveillance WHO protocol in Europe in late January^19^. In this regard, we performed backward simulations to estimate the day zero of the epidemic in Italy. We tested different combinations of initial undetected asymptomatic and symptomatic cases to determine the time needed to reach the values reported on February 24^th^. More details about the backward integration can be found in Methods. Our analysis suggests the presence of a few undetected cases in Italy already in the late November – early December, as assessed in a recent study that confirmed the presence of the virus in northern Italy wastewater on December 18^th^, 2019^20^. A similar conclusion was drawn from the presence of IgM/IgG antibodies against SARS-CoV-2 Nucleocapsid protein in blood samples collected in Milan at the start of the outbreak^21^.

Model predictions based on parameter estimations of late Phase I (April 14^th^ – May 4^th^) and late Phase II (May 18^th^ – June 15^th^) allowed us to compare the long-term epidemic curves for the two scenarios. In Figure 3, we observe that, unexpectedly, the simulated scenario with late Phase I parameters (full lockdown) predicts more death and infected cases, and less recovered cases than the late Phase II data and simulations. This computational evidence suggests that, while the end of the lockdown did not perceptibly increase the infection rates, the epidemic may be less virulent in terms of pathological effects. The use of the late Phase II data for the comparison reinforces this hypothesis since only the first weeks (May 4^th^-May 18^th^) are likely influenced by previous lockdown policies.

**Figure 3:**
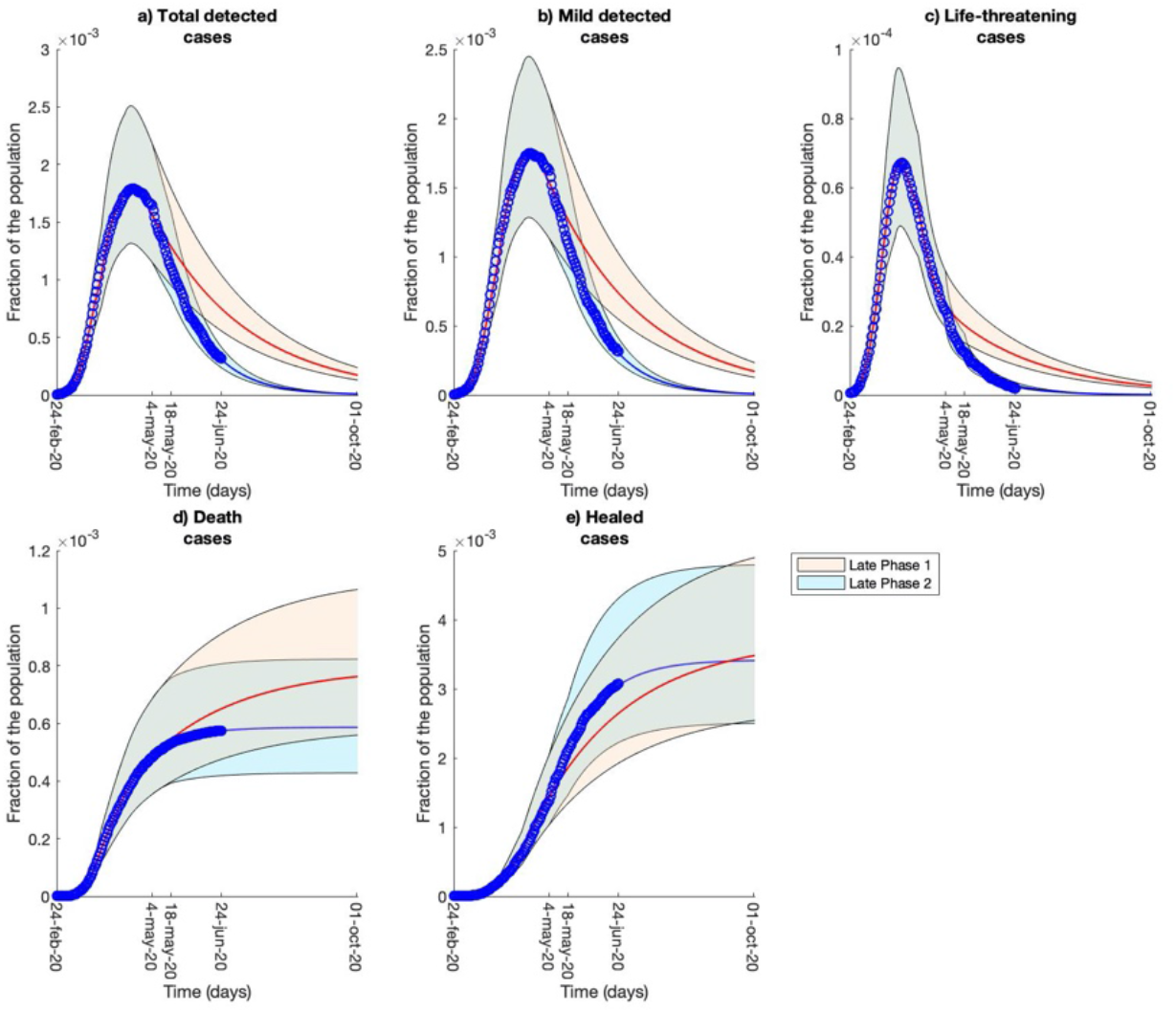
Comparison of model predictions. Comparison between long-term predictions based on the parameter estimates of late Phase I (from April 14th to May 4th, in red) and of late Phase II (from May 18th until June 24th, in blue). For each prediction, we reported the first, second and third quartiles. The blue dots represent the Italian data used for the calibrations.

Despite the natural reported evolution of the SARS-Cov-2 virus^22–25^, there are no clearcut genetic evidence of mutations that are weakening the virus. Thus, the virus may be losing strength in Italy due to external factors, such as weather conditions (temperature and humidity) and persistent consciousness and responsible behaviour (use of face masks, frequent hand washing, increased adoption of smart working), which may play a crucial role in the Italian COVID-19 evolution.

In the last part of our analysis, we focused on the evaluation of future scenarios. At this stage, it appears unlikely the eradication of COVID-19 only with social distancing measures and public health system efforts, as it was in 2003 for SARS-CoV-1^26^. On the contrary, SARS-CoV-2 may continue to circulate with a seasonal component, after the first epidemic wave, as it is for two different strains of human coronavirus, the HCoV-HKU1 and the HCoV-OC43^27,28^. Since at the moment no vaccines or preventative pharmaceutical treatments are readily available for COVID-19, we investigate the effects of different social distancing measures on possible future seasonal waves of COVID-19. Given the negative impact of the lockdown on the economy^29^, we analysed a trade-off between controlling a new epidemic wave and allowing the commercial activities and industries to be open for a continuative period.

We tested long-term scenarios considering a gradual worsening of the epidemic situation in the autumn/winter period. We implemented a seasonality function, defined on the basis of hyperbolic tangents, which smoothly changes the contagion parameters. We simulated an increase in the infection rates in the period October-November to reach a condition similar to the one experienced in early March 2020. On the contrary, they were decreased in April-May to restore the spreading condition of the current summer period. All the mathematical details can be found in Methods. In our simulations, we impose an intermittent lockdown^30^ (as opposed to the extended lockdown imposed during the first epidemic wave) and investigated the effect of an increasing number of “open days” (no lockdown) in each two-week period. The intermittent lockdown is introduced once the reported daily cases exceed 500 infected nation-wide for at least three consecutive days and is then simulated until the end of May. Indeed, starting from April, the spreading parameters are smoothly restored to the ones of the summer period (estimated in the subinterval May 18^th^ – June 24^th^) and the lockdown policy is no more necessary. Figure 4 shows the scenario where there are six “open days” every two weeks, while Extended Figure 3 reports the results for different combinations of “open days”.

**Figure 4:**
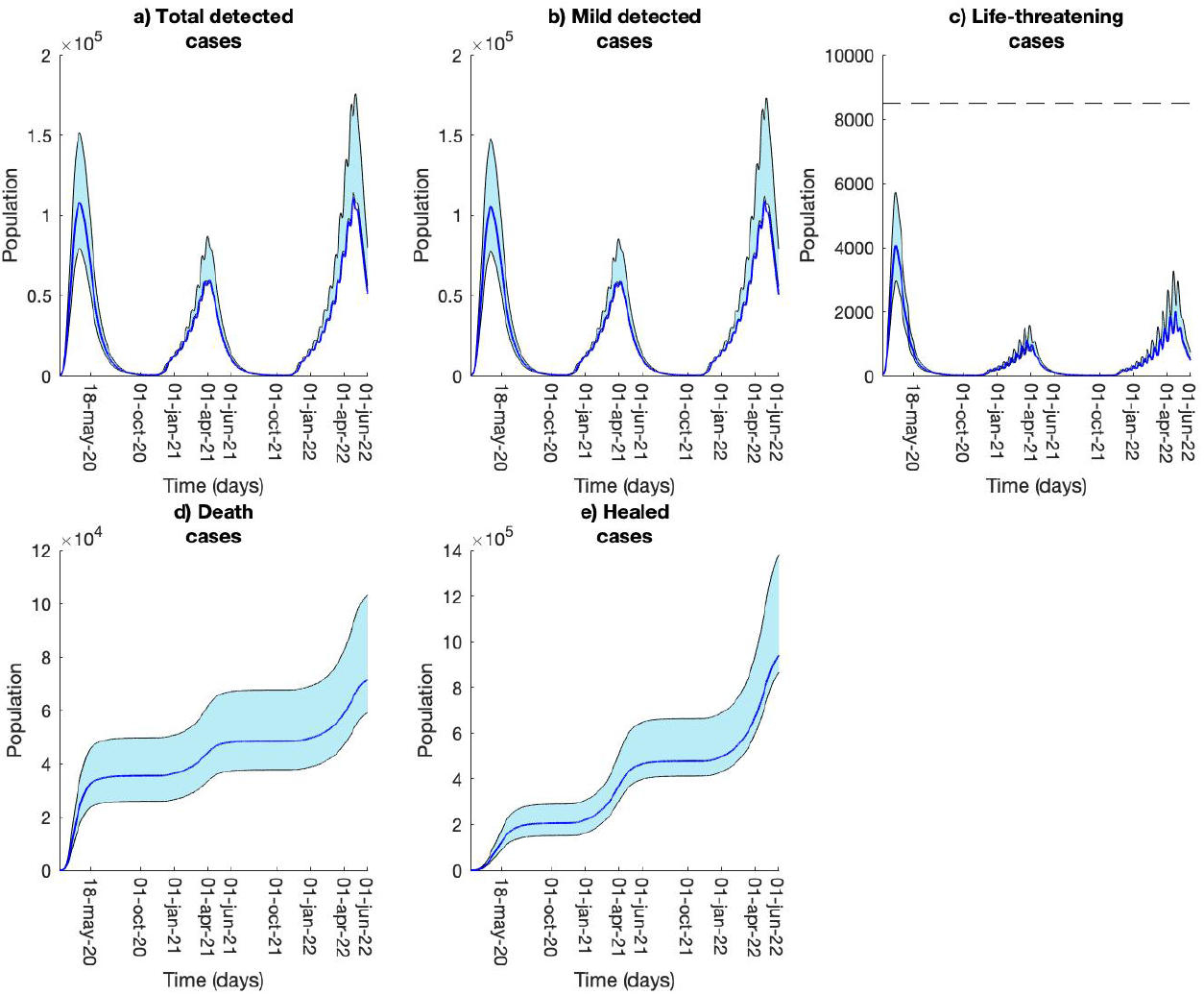
Long-term prediction with seasonal component and intermittent lockdown. Long-term model predictions computed by including a seasonal component to the contagion parameters and an intermittent lockdown policy. The lockdown alternates between 6 “open days” (no lockdown) and 8 “close days” (complete lockdown) when the epidemic situation worsens. The first peak refers to the first epidemic wave that we experienced in the period February-June 2020. The second and the third peaks are predictions for 2021 and 2022, respectively. The plots represent the first, second and third quartiles; the time length of the simulations goes from February 2020 until June 2022. The dashed horizontal line in panel c) is the maximum ICU capacity for Italy, reported as 8940 beds^31,32^.

Comparing the model simulations with the ICU-bed capacity, our results suggest that 6 over 14 days is the maximal threshold that can be endured by the Italian healthcare system. The prediction shows the median value for the epidemic curve, as well as the first and the third quartiles representing virtuous or bad habits of the Italian population, respectively. The second peak reaches its maximum value in April 2021, suggesting that the timely application of social distancing measures can delay the epidemic curve with respect to the 2020 peak. However, the delay gives rise to an increased number of infected cases during the summer, resulting in a 2022 epidemic wave that is higher than 2021.

A similar prediction is obtained when the intermittent lockdown is completely driven by daily infected cases. We considered the seasonal component, and we imposed the lockdown every time the daily number of infected exceeds 500 cases for at least three consecutive days, as in Figure 4. Once started, the lockdown is enforced without interruptions until the simulated daily number of infected decreases to less than 250 cases for at least one week. Extended Figure 4 shows that the median curve of the case-driven lockdown is slightly better than the one presented in Figure 4, suggesting that the number of infected people, in the long run, is lower. However, in a case-driven lockdown policy, the number of “open days” and “close days” (strict lockdown for the population) is not constant over the time, thus becoming harder to manage for the population. With the thresholds of 500 and 250 daily cases driving the lockdown, our results show a mean value, computed on the overall intermittent lockdowns, of 8 “open days” alternated with 12 “close days”, suggesting a slight advantage over the 6 “open days” and 8 “close days” in Figure 4. During the analysis, we also tested a scenario where only 150 daily cases were required for the opening policy, but no significant differences were observed in the model dynamics, suggesting that our model predictions seem to be poorly affected by the chosen opening threshold.

Our results suggest that, in the case of future seasonal outbreaks, an intermittent lockdown could be a valid alternative to the extensive lockdown enforced during the first epidemic wave. The intermittent strategy relieves the economic and psychological burdens on society, and is still able to contain the epidemic and avoid an exponential uncontrolled outbreak. Also, we had highlighted the importance of non-pharmaceutical interventions, such as social distancing measures, frequent hand washing and wearing protective masks. As we show in Figure 3, these precautions seem crucial to contain the epidemic spread, even without a lockdown. Hence, our study suggests that keeping correct behaviours represent the first line of defence against new COVID-19 outbreaks, both local and imported ones^33^.

The main limitation of these results is that all the long-term predictions may change due to the potential approval and distribution of new pharmaceutical interventions, either in forms of drugs or vaccines^34–36^. Also, our results represent the overall situation in Italy, without considering that each region had a very different first epidemic wave. To locally describe the epidemic, regional calibrations of the model as well as spatial interactions between the regions should be considered^6,37,38^. However, besides our analysis of the Italian situation as a case study, our time-varying protocol for model calibration is a general framework, and it may apply, with just minor efforts, to any available data on COVID-19. Another possible extension of this work could be the introduction of age classes^39,40^. COVID-19 has a different impact on the demographic tissue, with the younger population much less severely affected than the elderly; however, no public repository is currently available about daily age-structured cases, which would be needed for the model calibrations. Even considering these limitations, our analysis based on the SIDARTHE model provides insights into the dynamics of the spread during the various phases of the Italian first epidemic wave. Moreover, it allows us to test multiple scenarios informing the policymakers on possible strategies to contain resurgent outbreaks at a nation-wide scale.

## Data Availability

This works is based on publicly available data provided by the Italian Protezione Civile.

## EXTENDED FIGURES

**Extended Figure 1:**
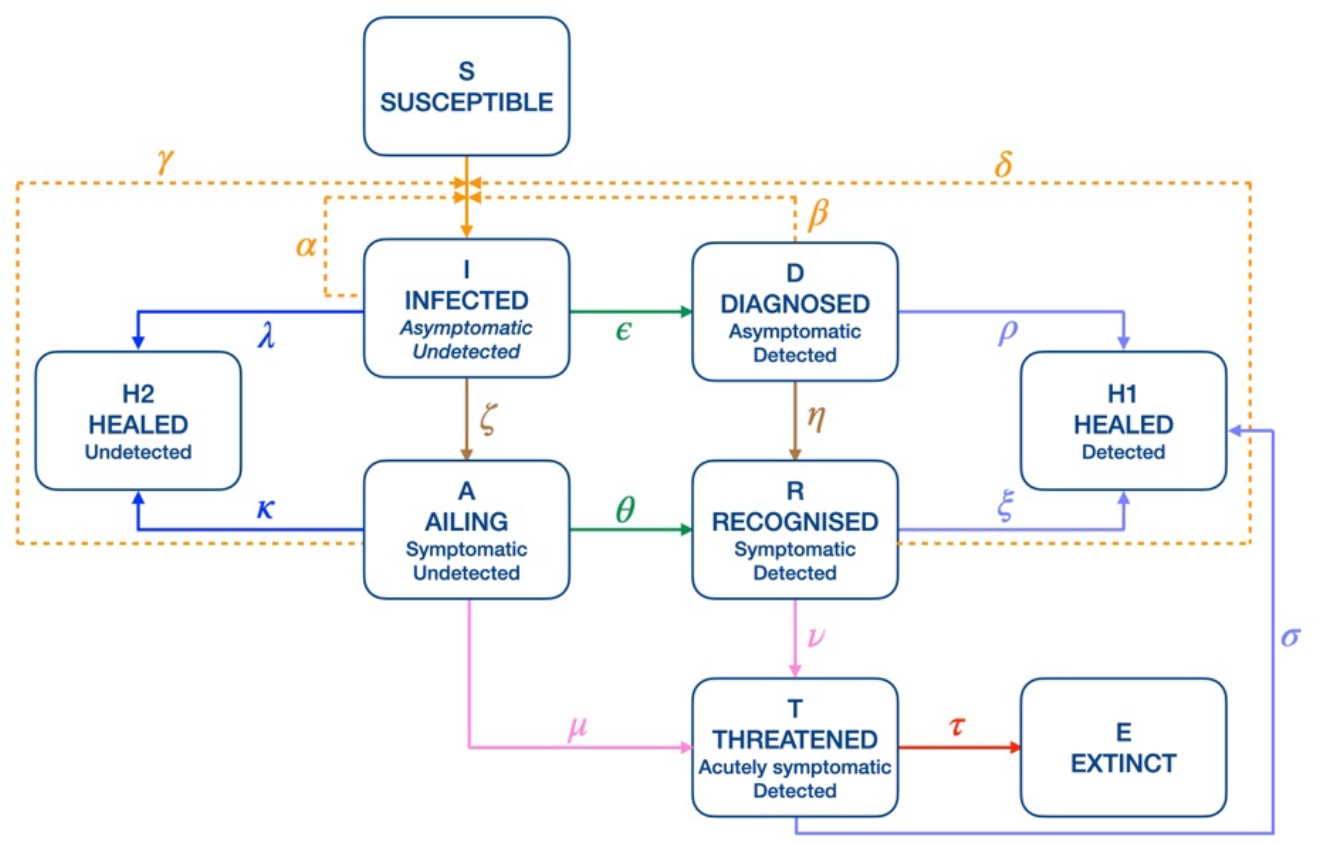
Network of the model. We considered the recently published SIDARTHE model^1^ and divided the healed class H between H1 and H2, representing the healed individuals from the detected and undetected classes, respectively. The continuous lines represent the movement of the individuals from one class to the other. The dashed lines represent the contagion of the susceptibles from one of the infected class. The color code of the lines is associated with the biological meanings of the parameters: orange for contagion, green for diagnosis, dark and light blue for healing, pink for the development of critical condition and red for death. The names of the model parameters are reported with Greek letters. Alpha: contagion rate due to asymptomatic undetected; Beta/Delta: contagion rates due to asymptomatic (beta) and symptomatic (delta) detected; Gamma: contagion rate due to symptomatic undetected; Epsilon: diagnosis rate for asymptomatic undetected; Lambda/Rho: healing rates for undetected (lambda) and detected (rho) asymptomatic; Theta: diagnosis rate for symptomatic undetected; Mu: rate of critical condition development for symptomatic undetected; Kappa/Xi: healing rates for undetected (kappa) and detected (xi) symptomatic; Nu: rate of critical condition development for symptomatic detected; Tau: death rate; Sigma: healing rate for ICU cases.

**Extended Figure 2:**
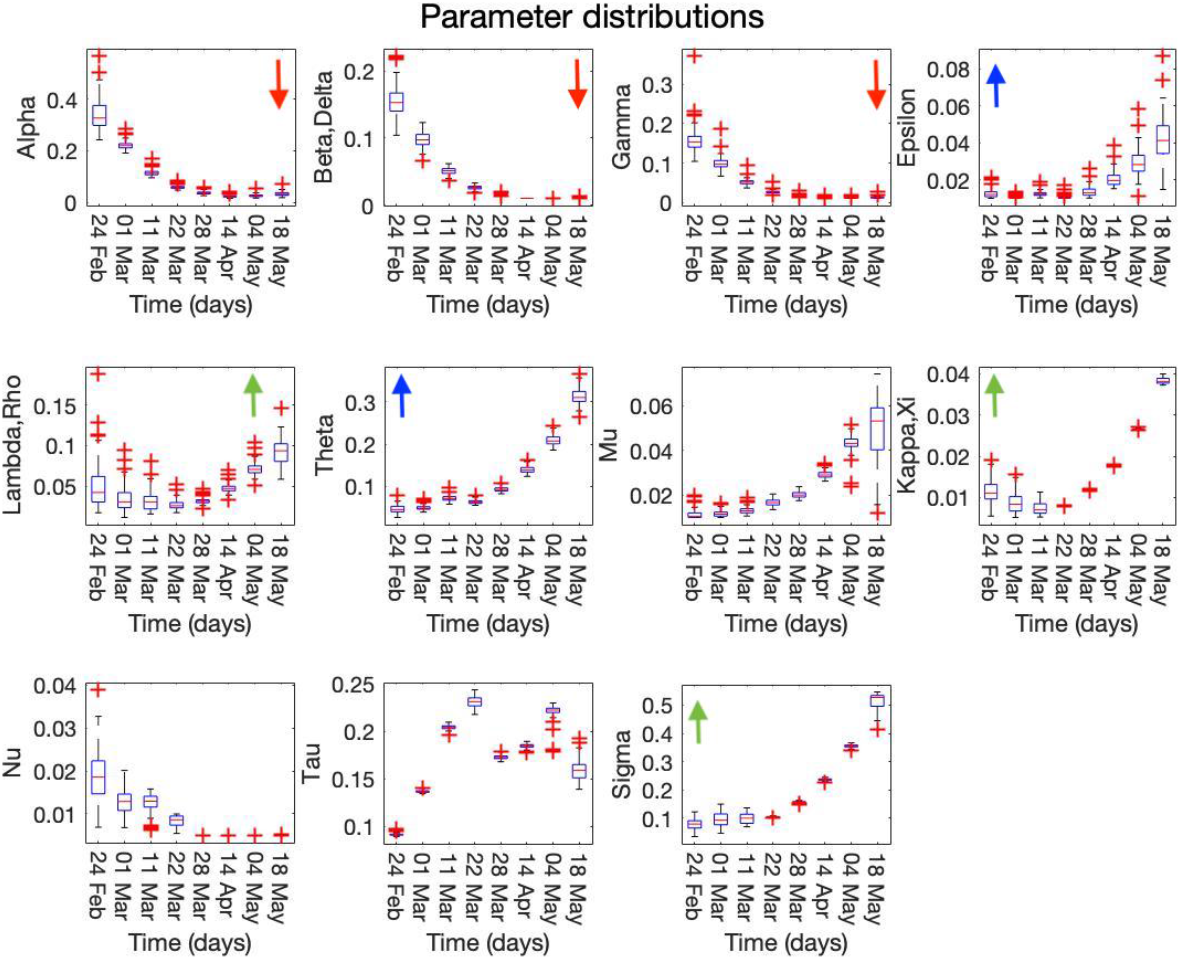
Parameter distributions. For each estimated parameter, we reported the distribution across the multiple calibrations at each critical date where the estimate value has been updated. We refer to the original SIDARTHE paper^1^ for a detailed discussion about the biological meaning of the parameters. We highlighted with red arrows the descending trend of the contagion parameters, with green arrows the ascending trend of the healing parameters and with blue arrows the ascending trend of the swab policies. Alpha: contagion rate due to asymptomatic undetected; Beta/Delta: contagion rates due to asymptomatic (beta) and symptomatic (delta) detected; Gamma: contagion rate due to symptomatic undetected; Epsilon: diagnosis rate for asymptomatic undetected; Lambda/Rho: healing rates for undetected (lambda) and detected (rho) asymptomatic; Theta: diagnosis rate for symptomatic undetected; Mu: rate of critical condition development for symptomatic undetected; Kappa/Xi: healing rates for undetected (kappa) and detected (xi) symptomatic; Nu: rate of critical condition development for symptomatic detected; Tau: death rate; Sigma: healing rate for ICU cases.

**Extended Figure 3:**
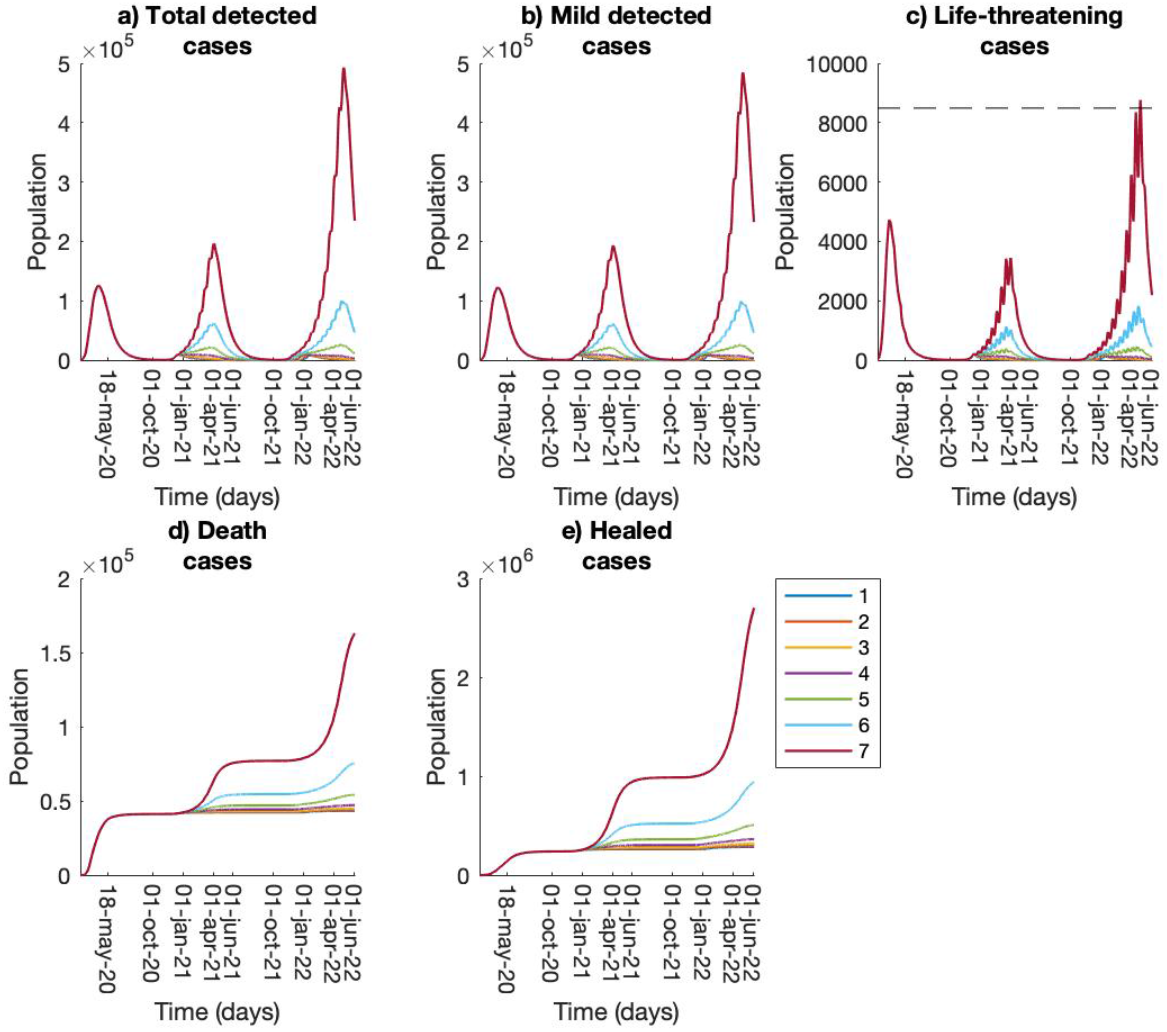
Sensitivity analysis of the number of open days. For the intermittent lockdown scenario, we tested different combinations of numbers of “close days” and “open days”. Here, we reported the model dynamics for 1 up to 7 “open days” (numbers in the legend) that are alternated with 13 up to 7 “close days” during the months from October to June. The simulations show that the scenario with 7 “open days” can be barely endured by the Italian healthcare system and almost reaches the maximum ICU-bed capacity (dashed horizontal line in panel c). Each dynamic evolution is computed with the median estimated parameter set.

**Extended Figure 4:**
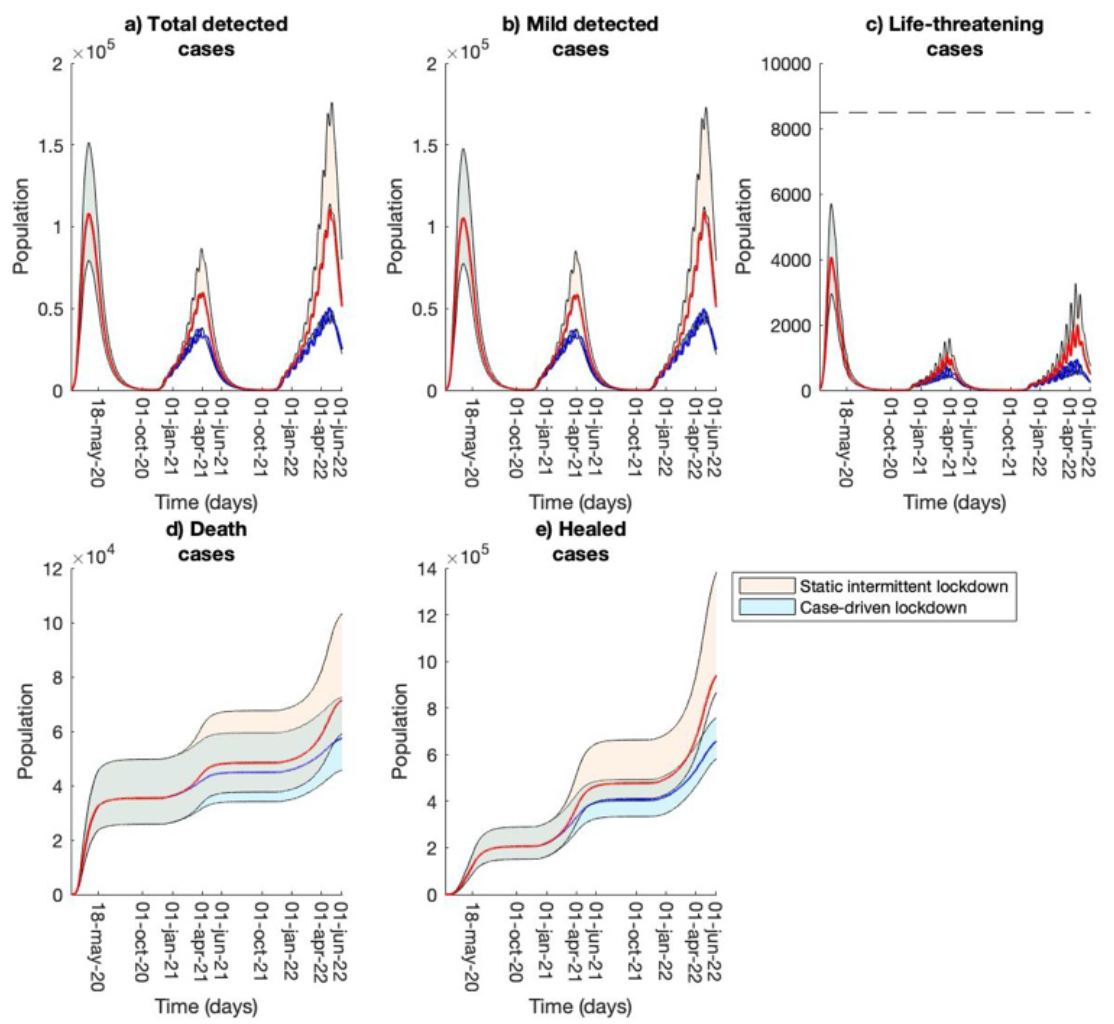
Long-term prediction with different intermittent lockdown polices. Comparison between long-term model predictions computed with different intermittent lockdown polices. The red curves represent an intermittent lockdown with a fixed number of 6 “open days” alternated with 8 “close days” during the months from October to June (scenario presented in Figure 4). The blue curves describe an intermittent lockdown that is driven by the number of daily infected cases. Specifically, the “close days” start when there are more than 500 daily new infections for at least 3 days. The “open days” start when the daily infections go below 250 cases for at least one week. During the period October - June, the lockdown alternates a mean of 8 “open days” with 12 “close days”. In both the two scenarios we included the seasonal component for the contagion parameters.

## METHODS

### The mathematical model

The original SIDARTHE model^1^ divides the entire population into 8 mutually exclusive compartments describing different infection stages: each individual can be either susceptible (S), undetected asymptomatic or pauci-symptomatic infected (I), detected asymptomatic infected (D), undetected symptomatic infected (A), detected symptomatic infected (R), detected life-threatened symptomatic infected (T), recovered (H) or dead (E). In this contribution we also included an explicit subdivision of the recovered patients between those who had been previously detected (H1) and those who had not (H2), which eases the parameter calibration for the model. Hence, the SIDARTHHE model we actually considered is a closed-form ODEs system involving 9 variables that closely retraces the original model:

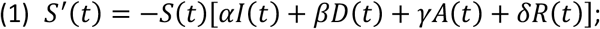

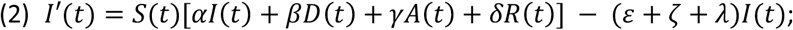

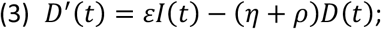

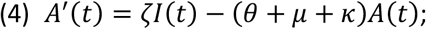

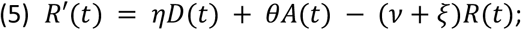

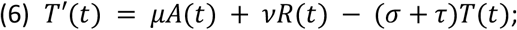

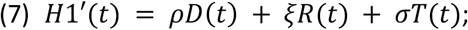

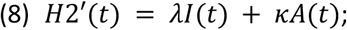

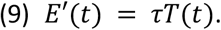

Extended Figure 1 visualizes the flows among different compartments, representing infection stages, which govern the epidemic evolution. We refer to the original work^1^ for more details on the model parameters.

### The time-varying calibration procedure

The data source used to estimate the model parameters is the GitHub page of the Italian Protezione Civile, where new data on the number of detected quarantined people, detected hospitalized patients, ICUs patients, recovered people (confirmed by two consecutive negative swabs) and deaths are loaded daily. We used data from February 24^th^, 2020 to June 24^th^, 2020, for a time period of 122 days.

We defined an *ad hoc* fitting strategy to manage the shifting policies occurred in the analyzed time interval by means of parameter updates reflecting those changes. The idea is that model parameters are not constant in time, but rather evolve in response to changes in policies and behaviors (*e*.*g*., washing hands more frequently and wearing face protective masks), and evolutions of the disease. In accordance with Italian national decrees and general measures, March 1^st^, 11^th^, 22^nd^, 28^th^, April 10^th^, May 4^th^ and 18^th^ have been selected as critical days to mark the parameter updates. In details, we estimated the model parameters for the first time subinterval (February 24^th^ – March 1^st^) and, for every subsequent subinterval (March 1^st^– March 11^th^, March 11^th^ - March 22^nd^, March 22^nd^ – March 28^th^, March 28^th^ – April 10^th^, April 10^th^ – May 4^th^, May 4^th^– May 18^th^ and May 18^th^ – June 24^th^) we estimated for each parameter a scaling factor (+/- 50%) that, once applied to the estimate of the previous subinterval, best fit the data of the corresponding time range. In this way, the value of the parameters in each subinterval is correlated with the corresponding value of the previous subintervals in a continuous way, thus reflecting a smoothly time-dependent adaptation of the parameter values. All parameter values and scaling factors were estimated within a single optimization procedure aiming at fitting the model to the complete time series. In this way, we reduced the risk of bias for the computed parameter estimates due to the length of some subintervals shorter than the COVID-19 incubation period. In addition, prior knowledge on standard epidemic mechanisms as well as on COVID-19-specificities has been used to constrain the objective function. In particular, we made assumptions in order to model the greater probability to be detected if symptoms are visible (parameter *θ* > parameter *ε*) and the relationships among infection rates, with undetected asymptomatic infected more likely to spread the epidemic than both undetected symptomatic (parameter *α* > parameter *γ*), which are supposed to stay at home to recover, and detected asymptomatic/symptomatic (parameter *α* > parameter *γ* > parameters *δ* and *β*), which are assumed to be quarantined. We assumed the same contagion rate for the two classes of detected cases (parameter *δ* = parameter *β*) and the same healing rate for the asymptomatic classes (parameter *λ* = parameter *ρ*) and the symptomatic ones (parameter *κ* = parameter *ξ*). We also imposed a maximum value for R0 equal to 6 as reported in the literature^41^. Moreover, we estimated the initial value of those model variables that do not have a direct corresponding value in the available data (I, D, A, R and H2) under the assumption that the number of undetected cases was greater than the detected one.

For the calibration procedure, we used a global optimization algorithm, the CMA-ES^7,8^, a derivative-free evolutionary strategy for optimization problems. The estimates for the model parameters are computed in the range 10^−2^ − 1 (except for *κ, v* and *ξ* for which the lower bound was fixed to 5. 10^−3^), while for the initial model variables we imposed a range equal to 10^−9^ − 10^−4^ (as fraction of the Italian population). The value for parameters *ζ* and *η* was fixed to 0.125, corresponding to a mean of 8 days to develop the symptoms.

We calibrated the model 100 times and worked with the resulting parameter distributions to compute the long-term predictions.

### Estimation of day zero of the epidemic

We estimated the actual day zero of the epidemic outbreak, when the first unrecognized cases occurred, with a “shooting” forward approach, which was preferred to the most common back integration technique because of the lighter computational burden. In particular, we selected an alleged, but reliable, number of undetected infected as initial conditions to be satisfied at day 0 and we used the estimated model parameters of the first subinterval (February 24^th^– March 1^st^) to simulate the model. We argued that the parameters of such a subinterval were the ones that best suit our purposes since they are not affected by any countermeasure and hence can better reflect the unawareness situation. The simulations start at day 0 and terminate when all the model variables are close to the values estimated at February 24^th^. The time interval needed to reach the termination criteria allowed us to find day zero on the calendar by going back from February 24^th^ for a number of days corresponding to the interval. Table 1 shows some results we obtained by varying the initial conditions fulfilled at day zero. Other combinations for the initial conditions, involving at most 10 patients, were considered to check the consistency of the estimations. All of them predicted the day 0 to lie between late November and early December.

**Table 1:** Different combinations of asymptomatic and symptomatic undetected cases were tested to predict the day 0 of the epidemic outbreak. Each parameter set computed with the repeated calibrations has been used to make the prediction, thus we reported for day 0 the mean and standard deviation of the results. Also, we included minimum and maximum of the distribution results. For the simulations, all the model variables, except for I and A, were set to zero.

| Supposed initial conditions at day 0 | Predicted day 0 (mean +/- std) |
| --- | --- |
| I (asymptomatic undetected) = 0;<br>A (symptomatic undetected) = 1 | 20 November +/- 4 days<br>(min: 10 November, max: 1 December) |
| I = 1;<br>A = 0 | 21 November +/- 4 days<br>(min: 11 November, max: 2 December) |
| I = 1;<br>A = 1 | 21 November +/- 4 days<br>(min: 11 November, max: 3 December) |
| I = 2;<br>A = 2 | 23 November +/- 4 days<br>(min: 14 November, max: 5 December) |
| I = 10;<br>A = 1 | 30 November +/- 4 days<br>(min: 20 November, max: 10 December) |

### The seasonal component in the long-term predictions

In all the long-term predictions we considered the first, second and third quartiles computed from the parameter distributions to represent an optimistic, median and pessimistic scenario, respectively.

We imposed a seasonal component for the contagion parameters in the long-term predictions. The four contagion rates (*α, β, γ* and *δ*) are defined as:

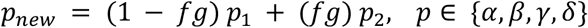

where *p*_1_ is the estimate value for the contagion parameter in the last subinterval (May 18^th^ – June 24^th^), *p*_2_ is the mean value of the contagion estimates in the first two subintervals (February 24^th^ – March 1^st^ and March 1st– March 11st) when the lockdown was not yet active, *f* is defined as a hyperbolic tangent that mimics a smooth increase of the contagion parameters during a time interval of two months (October-November), 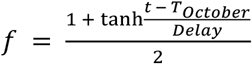, and *g* is defined as a hyperbolic tangent that mimics a smooth decrease of the contagion parameters during a time interval of two months (April-May), 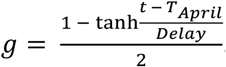. For both the hyperbolic tangents, the *Delay* is equal to two months while *T*_*october*_ and *T*_*April*_ correspond to the first of October and April, respectively. With this procedure, we were able to simulate a smooth increase of the contagion parameters starting at October 1 that requires around two months to grow from the smaller value *p*_1_ to the greater value *p*_2_. On the contrary, we have a smooth decrease that starts on April 1^st^ and requires around two months to decrease from the greater value *p*_2_ to the smaller value *p*_1_.

### Intermittent lockdown in the long-term predictions

We implemented an intermittent lockdown^30^ taking place during the seasonal outbreak of the epidemic (from October to June). The lockdown starts when the number of daily infected cases is above 500 for at least three consecutive days. During the period of lockdown, we alternate “close days” where a full lockdown is imposed to the population and “open days” where no lockdown is present. To simulate the alternation between close and open days, the contagion parameters are changed between the values *p*_1_ and *p*_2_, respectively, as detailed in the previous section. However, in this case, parameters *p*_2_, representing the contagion rates during the “close days” of the lockdown, are set equal to the mean of the contagion estimates during the last two subintervals of the extended lockdown (March 28^th^ – April 14^th^ and April 14^th^ – May 4^th^).

In our analysis, two different scenarios were tested for the intermittent lockdown. In the first scenario, we imposed a constant amount of open and close days and we tested different combinations of the two. In the second scenario, the lockdown is completely driven by the number of daily infected cases. A close policy starts every time there are more than 500 detected cases for at least three days. On the contrary, an open policy starts when there are less than 250 daily cases for at least one week.

